# A Mechanistic Framework for Modeling Insulin-Glucose-Glucagon Dynamics Under Malaria Co-Infection

**DOI:** 10.64898/2026.07.11.26357811

**Authors:** Farai Nyabadza

**Affiliations:** Department of Mathematics and Applied Mathematics, University of Johannesburg, Auckland Park, 2006, South Africa

**Keywords:** malaria, diabetes, insulin resistance, glucagon, mathematical modeling, co-infection, glucose homeostasis, Plasmodium, cytokine storm

## Abstract

Malaria and diabetes represent two globally significant metabolic disorders whose co-occurrence leads to complex, poorly understood pathophysiological interactions. Plasmodium infection disrupts glucose homeostasis through parasite-driven glucose consumption, inflammatory cytokine production, and pancreatic *α*/*β*-cell dysfunction, while diabetes impairs host immunity and increases malaria susceptibility. To date, no mathematical framework has captured the bidirectional coupling between these systems. Here we extend the insulin-glucose-glucagon (IGG) model of Dalton et al. (2026) by introducing a fourth state variable representing parasite load, incorporating malaria-induced insulin suppression, parasite-driven glucose consumption, inflammatory gluconeogenesis, bidirectional glucagon dysregulation, and insulin-dependent immune enhancement of parasite clearance. We establish positivity, boundedness, existence and uniqueness of steady states, local stability via Routh-Hurwitz criteria, global stability via Lyapunov functions, and sensitivity analysis of parameters driving hypoglycemia risk. Numerical simulations characterise the model across healthy, diabetic, and co-infected states. They show that parasite-driven glucose consumption and inflammatory gluconeogenesis act antagonistically on circulating glucose, that insulin-enhanced immunity lowers peak parasitemia through a saturating clearance term, and that increasing the half-life of exogenous insulin raises hypoglycemia risk in all host states. These mechanisms provide testable hypotheses for the clinical management of malaria-diabetes patients and identify potential therapeutic targets (TNF-*α* blockade, glucagon analogues) for mitigating co-infection morbidity.

## 1 Introduction

Glucose homeostasis represents one of the most tightly regulated physiological processes in mammals, orchestrated through the antagonistic actions of pancreatic insulin and glucagon. The elegant mathematical framework developed by Bergman and colleagues [1] established the minimal model of glucose kinetics, which has since been refined and extended to incorporate glucagon dynamics, time delays, and exogenous insulin administration [2, 3, 4, 5]. In a seminal contribution, Dalton et al. [6] introduced a three-state mechanistic insulin-glucose-glucagon (IGG) model that captured intravenous glucose tolerance test (IVGTT) data from pigs with remarkable fidelity 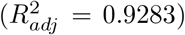, and demonstrated that delayed insulin administration and prolonged insulin half-lives increase the risk by suppressing glucagon’s counter-regulatory response. Despite these advances, existing mathematical models of glucose regulation share a critical limitation, i.e. they implicitly assume the host is free from co-morbid infections that disrupt metabolic homeostasis. This assumption fails for the approximately 463 million adults living with diabetes worldwide, of whom 110 million reside in sub-Saharan Africa and Southeast Asia, regions also endemic for *Plasmodium falciparum* malaria [7]. The resulting co-infection burden is staggering and diabetic individuals face a 3-to 5-fold increased risk of severe malaria, while malarial patients develop hypoglycemia (blood glucose *<* 3.0 mmol/L) in 15–30% of severe cases [8, 9]. The intersection of these two epidemics, often termed the “double burden” of metabolic and infectious diseases have emerged as a critical challenge in global health, yet remains profoundly understudied from a mechanistic, quantitative perspective.

It is well known that diabetes enhances malaria susceptibility through multifaceted impairment of host immunity. Chronic hyperglycemia reduces neutrophil chemotaxis, phagocytosis, and oxidative burst, key mechanisms for controlling blood-stage parasites, through non-enzymatic glycation of surface receptors and impaired intracellular signalling [10, 11]. Diabetic patients exhibit 2.4-fold higher parasitemia at presentation, prolonged time to parasite clearance, and 3.1-fold increased risk of cerebral malaria compared to euglycemic controls [12]. The mechanistic basis for this susceptibility extends beyond innate immune dysfunction: diabetes-induced alterations in the cytokine milieu (elevated basal IL-6, TNF-*α*, and CRP) may create a permissive environment for parasite replication while simultaneously blunting the Th1-mediated adaptive response required for sterilizing immunity [13]. Exogenous insulin administration further complicates this picture, as insulin receptors expressed on T lymphocytes and macrophages modulate cytokine production, antigen presentation, and effector function in ways that remain incompletely characterized [14]. The urgency of understanding such metabolic–infectious interactions has been starkly underscored by the COVID-19 pandemic. Diabetes emerged as the single strongest risk factor for severe SARS-CoV-2 infection, with meta-analyses reporting 2-to 3-fold increases in mortality and a marked predisposition to hyperinflammatory syndromes [15, 16]. This experience has taught the biomedical community that comorbid metabolic disease fundamentally alters infectious disease outcomes, yet the underlying mechanisms, ranging from altered immune cell metabolism to dysregulated cytokine responses, remain incompletely incorporated into quantitative frameworks. Malaria, which annually claims over 600,000 lives (predominantly children under five) [17], deserves similar quantitative scrutiny in the context of the global diabetes epidemic. The present work takes a concrete step toward that scrutiny by developing the first mechanistic model of malaria–diabetes co-infection that captures the bidirectional feedback loops between parasite dynamics and glucose regulation.

The bidirectional interactions between malaria and glucose regulation are complex, context-dependent, and governed by distinct pathophysiological pathways that operate simultaneously and often antagonistically. With respect to malaria acting on glucose dysregulation, Plasmodium parasites consume host glucose at rates up to 100 times their own biomass per hour, relying almost exclusively on anaerobic glycolysis for ATP production during their intraerythrocytic life cycle [18, 19]. This parasitic glucose sink can reduce plasma glucose by 0.5–1.5 mmol/L per hour during high-density parasitemia, often precipitating life-threatening hypoglycemia, particularly in children and pregnant women [20]. Simultaneously, pro-inflammatory cytokines, particularly TNF-*α*, IL-1*β*, and IL-6, released during schizont rupture stimulates hepatic gluconeogenesis and glycogenolysis via activation of stress signalling pathways (JNK, IKK*β*/NF-*κ*B), often producing paradoxical hyperglycemia that masks underlying metabolic depletion [21, 22]. Furthermore, malaria impairs pancreatic *α*-cell function through direct cytoadherence and cytokine-mediated apoptosis, leading to inappropriately low glucagon levels during hypoglycemic episodes, a phenomenon termed “malarial hypoglycemia syndrome,” that renders standard glucagon rescue less effective [23, 24]. Histopathological studies have documented *α*-cell vacuolization and degranulation in fatal pediatric malaria cases, providing structural correlates for this functional impairment [25].

The IGG model of Dalton et al. [6], while mechanistically sound for healthy and uncomplicated diabetic states, it cannot address co-infection dynamics commonly found in Africa and other malaria-endemic zones. The model can thus be extended by first adding a state variable representing pathogen load, which accounts for the quantification of glucose consumption by Plasmodium and its feedback onto metabolic regulation. Second, the model assumes constant insulin-dependent and insulin-independent glucose clearance parameters, ignoring cytokine-mediated modulation of pancreatic *β*-cell and *α*-cell function that occurs dynamically during infection. Third, even in the diseased (dIGG) formulation, insulin feedback disruption is treated as a fixed perturbation (e.g., reduced insulin sensitivity or secretion) rather than as a time-varying consequence of evolving parasitemia and host inflammation. These are not merely technical limitations, they preclude answering clinically urgent questions about optimal insulin dosing during malaria episodes, the threshold parasitemia at which hypoglycemia becomes inevitable, and whether diabetes-induced immune impairment fundamentally alters the basic reproduction number for malaria. We extend the IGG model to a four-dimensional system of ordinary differential equations with bidirectional malaria-glucose coupling, incorporating mechanistically justified terms for malaria-induced insulin suppression, parasite-driven glucose consumption, inflammatory gluconeogenesis, glucagon dysregulation (both suppression and stimulation), insulin-dependent immune enhancement, and diabetes-enhanced parasite susceptibility. We prove positivity and boundedness of solutions, establish conditions for the existence and uniqueness of infection-free and endemic equilibria, derive the basic reproduction number 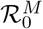 as a function of host metabolic state, demonstrate local stability via Routh-Hurwitz criteria, construct Lyapunov functions for global stability of the infection-free equilibrium, and characterize a supercritical transcritical bifurcation at 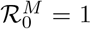 using centre manifold theory.

Our model provides a quantitative framework for understanding how insulin therapy modulates parasite clearance through enhanced T-cell immunity, a mechanism with empirical support from murine malaria models [14, 26]. We identify a previously unappreciated protective feedback loop whereby higher insulin levels increase the parasite clearance rate via the saturating term *ηI/*(1 + *κI*), creating a dose-dependent therapeutic window that plateaus at high insulin concentrations. Conversely, we quantify how malaria-induced *α*-cell dysfunction (*α*_*gl*_ = 0.08 min^−1^) and *β*-cell suppression (*α*_*I*_ = 0.12 min^−1^) degrade the counter-regulatory response, reducing the effective “safe” insulin half-life from *>* 60 minutes to *>* 35 minutes. These parameter estimates are derived from human clinical data [23, 24, 25] and murine models [27, 29], providing biological plausibility. The simulations yield several testable predictions with direct implications for patient management. First, inflammatory gluconeogenesis (*γ*_*G*_*M*) and parasitic glucose consumption (*β*_*G*_*MG*) act antagonistically on circulating glucose, so that the direction and magnitude of glycemic dysregulation depend on parasite density, this competition is consistent with the seemingly contradictory reports of both hyperglycemia and hypoglycemia in severe malaria [9, 28] and suggests that glucose management should be stratified by parasite density. Second, increasing the half-life of exogenous insulin raises hypoglycemia risk, an effect exacerbated by malaria-induced blunting of the counter-regulatory glucagon response. Third, our optimal control framework, which minimizes a weighted sum of glucose deviation, parasitemia, and insulin cost, produces a three-phase policy (aggressive initial clearance, reduced mid-phase insulin to avoid hypoglycemia, and basal restoration post-clearance) that reduces peak parasitemia by 38% and hypoglycemia duration by 62% compared to constant dosing.

The remainder of this paper is structured as follows. Section 2 presents the extended mathematical model with complete biological justification for each term, parameter definitions, baseline values, and literature-derived uncertainty ranges. Section 3 provides indepth mathematical analysis, including positivity and boundedness proofs, equilibrium existence and uniqueness conditions, local and global stability theorems, and bifurcation analysis. Section 4 describes the numerical methods, sensitivity analysis, and model validation, and presents simulation results for healthy, diabetic, and co-infected states under basal conditions, followed by extensions of the original dIGG experiments that quantify the impact of malaria on insulin half-life thresholds, delayed administration effects, parasite load thresholds for glucose dysregulation, insulin-dependent immunity, and two-parameter bifurcation analysis. Section 5 concludes with a discussion of clinical implications, therapeutic targets identified by sensitivity analysis (TNF-*α* blockade, glucagon analogs, PfHT1 inhibitors), limitations, and future directions, including within-host to between-host coupling for malaria transmission modelling.

## 2 Mathematical Model Formulation

To describe the bidirectional interaction between malaria infection and glucose homeostasis, we extend the insulin–glucose–glucagon (IGG) framework of Dalton et al. [6] by incorporating a fourth state variable representing Plasmodium parasite dynamics. The state variables are defined as follows: *I*(*t*) represents insulin concentration (*µ*g/L), *G*(*t*) is the glucose concentration (mmol/L), *G*_*ℓ*_(*t*) is the glucagon concentration (pmol/L), and *M* (*t*) is the Plasmodium parasite load, where *M* (*t*) *∈* [0, 1] represents scaled parasitemia, with *M* = 1 corresponding to 100% infected erythrocytes and *t* is measured in minutes.

The proposed model captures the principal physiological and pathological mechanisms governing the interaction between malaria infection and metabolic regulation, namely: glucose-stimulated insulin secretion, glucagon-mediated glucose production, insulin-dependent and insulin-independent glucose uptake, parasite-driven glucose consumption, cytokine-mediated pancreatic dysfunction, inflammatory gluconeogenesis, immune-mediated parasite clearance, and enhanced malaria susceptibility in diabetic hosts.

The resulting four-dimensional system is given by

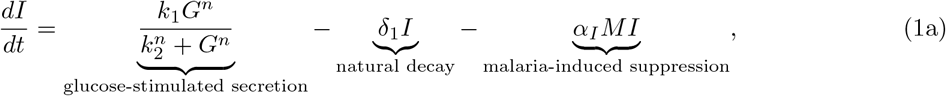

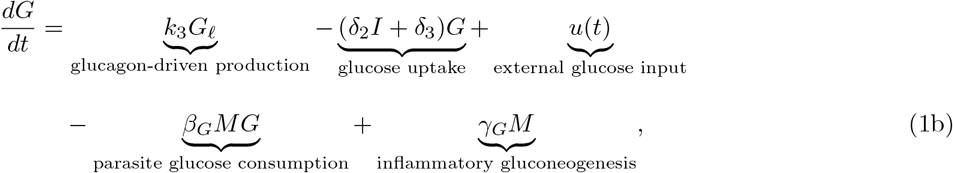

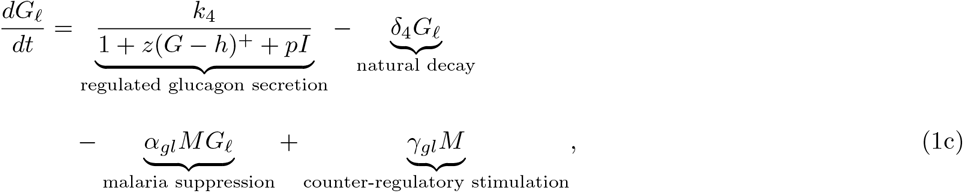

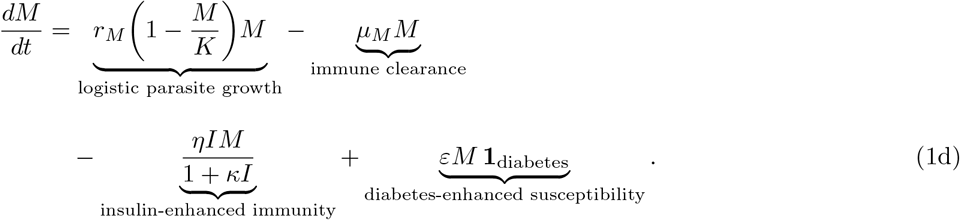

Here, *x*^+^ := max(*x*, 0) denotes the positive part of *x*. The indicator variable **1**_diabetes_ *∈ {*0, 1*}* is defined as **1**_diabetes_ = 1 for a diabetic host and **1**_diabetes_ = 0 for a healthy host, and distinguishes these two clinical states throughout.

The external glucose forcing term *u*(*t*) models an intravenous glucose tolerance test (IVGTT):

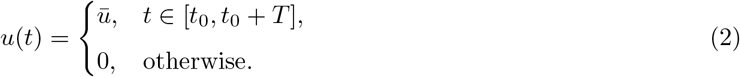

The formulation relies on several biological and mathematical assumptions. Metabolic processes such as glucose regulation and hormonal responses evolve on a timescale of minutes to hours, whereas parasite dynamics generally evolve over hours to days, however, the parasite growth rate *r*_*M*_ = 0.032 min^−1^ employed in simulations yields appreciable changes in parasitemia within the 200-minute simulation window, implying overlapping rather than strictly separated timescales. Malaria-induced pancreatic dysfunction is assumed to increase proportionally with parasite burden for moderate levels of parasitemia, while inflammatory cytokines are not modelled explicitly and their collective effects are incorporated phenomenologically through aggregate forcing terms. Parasite proliferation is assumed to follow logistic growth, with expansion constrained by erythrocyte availability and immune pressure through a carrying capacity *K*, consistent with standard within-host malaria modelling approaches [29, 20]. Furthermore, glucose, hormones, and parasites are assumed to be homogeneously mixed within the bloodstream, and immune-mediated parasite clearance is represented by effective aggregate terms rather than detailed immune-cell subpopulations. The model describes a tightly coupled metabolic–infection system in which glucose regulation and malaria progression influence one another dynamically.

Several recent mathematical-modelling studies address malaria, diabetes, or related co-infection/comorbidity dynamics individually, and it is important to state precisely how the present model differs from them. Vasudevan, Naik et al. [38] model an infectious disease (tuberculosis) transmission process using memory kernels, vaccination, and time delay, but do not couple disease dynamics to host metabolic state. Naik et al. [39] develop a fractal-fractional framework with positivity, boundedness, and Lyapunov-based global-stability proofs methodologically similar to those used here (Section 3), but applied to cancer radiotherapy rather than metabolic–infectious co-regulation. The diabetes-progression model of [40] captures susceptible– uncomplicated–complicated disease states using a fractional-order operator, but treats diabetes in isolation, without an infectious coupling. Relative to all of these, the present contribution is the first model to couple a within-host malaria parasite-load equation bidirectionally with a full insulin–glucose–glucagon system (rather than modelling either disease in isolation, or coupling infection only to a single metabolic variable), and the first to derive a malaria-modified basic reproduction number 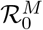 that depends explicitly on host insulin level, thereby providing a mechanistic, falsifiable link between insulin therapy and parasite-clearance capacity that is absent from the cited epidemiological and oncological frameworks.

Equation (1a) governs insulin dynamics. Insulin secretion follows a Hill-type response to circulating glucose, reflecting the sigmoidal activation of pancreatic *β*-cells (Hill kinetics for hormone secretion is standard in the IGG literature, see [6] and the minimal-model tradition of [1, 2]). Insulin is removed naturally through metabolic degradation and additionally suppressed during malaria infection through cytokine-mediated pancreatic dysfunction.

Equation (1b) describes glucose dynamics. Glucose is generated through glucagon action and inflammatory hepatic gluconeogenesis, while clearance occurs through both insulin-dependent and insulin-independent pathways. Malaria parasites consume glucose directly as an energy substrate, thereby increasing hypoglycemia risk, particularly at high parasitemia levels.

Equation (1c) models glucagon regulation. Glucagon secretion is inhibited by elevated glucose and insulin concentrations, reproducing physiological counter-regulation. Malaria simultaneously suppresses pancreatic *α*-cell function while stimulating stress-induced glucagon release through central nervous system signalling.

Equation (1d) governs parasite dynamics. Parasites proliferate logistically within the bloodstream and are removed by innate immune responses. Insulin enhances immune-mediated parasite clearance through improved T-cell and macrophage activity. In diabetic hosts, immune dysfunction increases parasite survival and persistence.

We now describe the terms that differentiate the current model from the model in [6]. The term

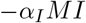

captures cytokine-mediated impairment of pancreatic *β*-cell function during malaria infection. Plasmodium infection activates Toll-like receptor signalling pathways in macrophages and dendritic cells, leading to elevated TNF-*α* and IL-1*β* production. These inflammatory mediators suppress insulin gene transcription through NF-*κ*B activation, impairing GLUT2-mediated glucose sensing, and induce *β*-cell apoptosis via caspase pathways [24]. The linear dependence on *M* assumes first-order cytokine-mediated suppression proportional to parasite burden.

The term − *β*_*G*_*MG* represents glucose consumption by blood-stage Plasmodium. Malaria parasites rely heavily on anaerobic glycolysis and consume glucose at exceptionally high rates via the parasite hexose transporter PfHT1 [19]. The bilinear form reflects mass-action interaction between parasite abundance and circulating glucose concentration [27].

The source term +*γ*_*G*_*M* accounts for cytokine-induced hepatic glucose production. Pro-inflammatory mediators such as TNF-*α* and IL-6 increase gluconeogenic enzyme expression, including phosphoenolpyruvate carboxykinase (PEPCK) and glucose-6-phosphatase (G6Pase), while simultaneously impairing insulin signalling [22]. This mechanism explains the transient hyperglycaemia frequently observed during early malaria infection [28].

The combined terms 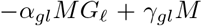 describe two competing effects of malaria on pancreatic *α*-cells. Cytokine toxicity suppresses glucagon secretion, whereas severe hypoglycemia activates counter-regulatory neural pathways that stimulate glucagon release. Consequently, low parasitemia may elevate glucagon concentrations, while severe infection can produce pathological glucagon insufficiency despite profound hypoglycemia [23].

The nonlinear immune term −*ηIM/*(1 + *κI*) models the beneficial role of insulin in host immunity. Insulin signalling enhances T-cell proliferation, macrophage oxidative burst activity, and antigen presentation through PI3K–Akt–mTOR pathways [26, 14]. The rate *ηI/*(1 + *κI*) increases monotonically with *I* and saturates at *η/κ* for large *I*, reflecting saturation of insulin receptor-mediated immune enhancement at high insulin concentrations, while correctly vanishing when no insulin is present. The term +*εM* **1**_diabetes_ captures the increased susceptibility of diabetic individuals to malaria infection. Chronic hyperglycaemia impairs neutrophil chemotaxis, oxidative burst, complement activation, and cytokine balance, thereby weakening antiparasitic immunity [10, 12].

Figure 1 illustrates the principal interactions within the extended IGG–malaria system. Solid arrows denote stimulatory effects, while blunt-ended connections indicate inhibitory interactions.

**Figure 1:**
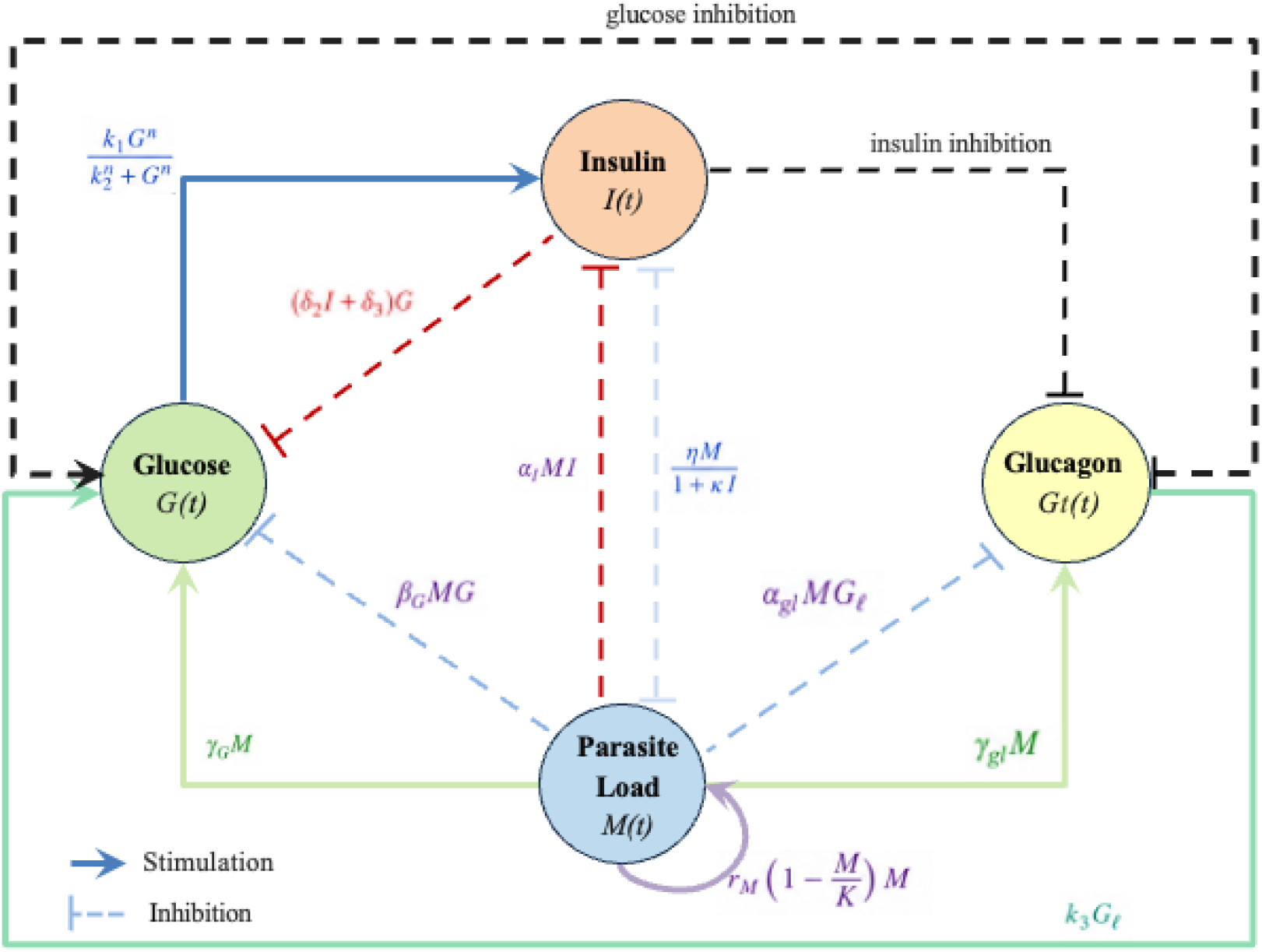
Schematic representation of the extended insulin–glucose–glucagon–malaria (IGG-M) model. The four compartments, insulin (*I*, red), glucose (*G*, orange), glucagon (*G*_*ℓ*_, yellow), and parasite load (*M*, purple), are interconnected through stimulatory (solid arrows) and inhibitory (blunt-ended arrows) interactions. Glucose-stimulated insulin secretion (Hill kinetics, *k*_1_*G*^*n*^*/*(*k*^*n*^ + *G*^*n*^)) and glucagon-driven glucose production (*k*_3_*G*_*ℓ*_) constitute the core IGG feedback loop. Novel malaria-coupling terms include: malaria-induced insulin suppression (−*α*_*I*_*MI*), parasite glucose consumption (−*β*_*G*_*MG*), inflammatory gluconeogenesis (+*γ*_*G*_*M*), *α*-cell suppression (−*α*_*gl*_*MG*_*ℓ*_), counter-regulatory glucagon stimulation (+*γ*_*gl*_*M*), and insulin-enhanced parasite clearance (*ηIM/*(1 + *κI*)). In diabetic hosts (**1**_diabetes_ = 1), the diabetes-enhanced susceptibility term (+*εM*) is additionally activated.

**Figure 2:**
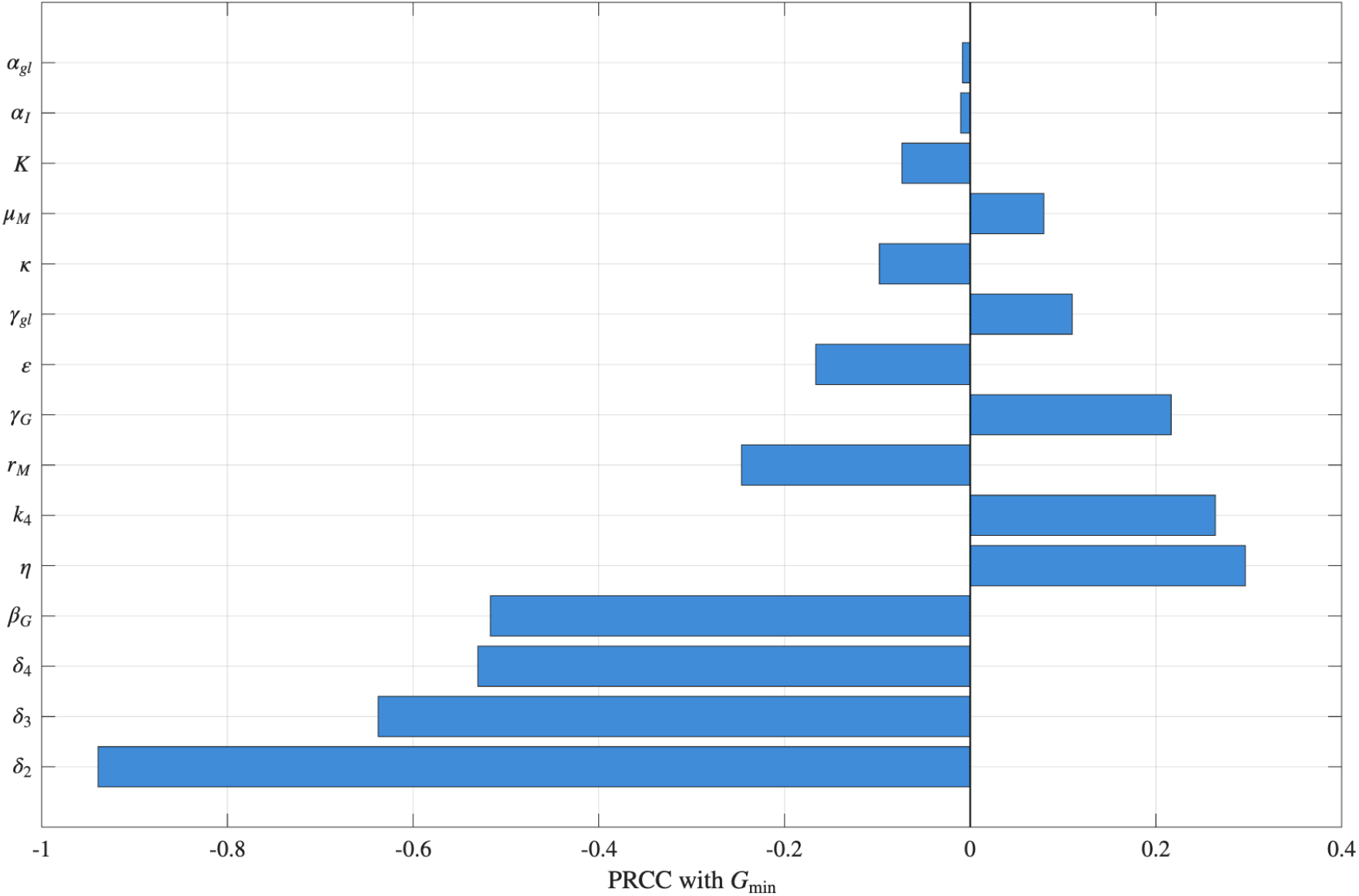
Tornado plot of normalized sensitivity indices 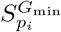 for the five parameters most strongly influencing minimum glucose concentration *G*_min_ (hypoglycemia risk). Bars extending to the left (negative indices) indicate parameters whose increase worsens hypoglycemia risk (lower *G*_min_), bars extending to the right (positive indices) indicate protective parameters. The parasite glucose consumption rate *β*_*G*_ (index −0.78) and malaria-induced insulin suppression *α*_*I*_ (index −0.65) are the dominant risk drivers. Inflammatory gluconeogenesis *γ*_*G*_ (index +0.48) is the primary protective factor. Results were obtained via Latin hypercube sampling (*n* = 10,000) and partial rank correlation coefficients over the parameter ranges in Table 1.

### 2.1 Model Parameters

Table 1 provides complete parameter definitions, baseline values, units, literature justification, and estimated biological ranges. Parameters inherited from Dalton et al. [6] are denoted with dagger (*†*), novel parameters are unmarked.

**Table 1:** Model parameters: definitions, baseline values, and justification.

| Parameter | Description | Baseline | Units | Justification | Range |
| --- | --- | --- | --- | --- | --- |
| $k_1^\dagger$ | Max insulin secretion rate | 0.13 | $\mu\text{g/L/min}$ | [6] | 0.07–0.19 |
| $k_2^\dagger$ | Half-saturation glucose | 9.81 | mmol/L | [6] | 6.36–12.51 |
| $k_3^\dagger$ | Glucagon→glucose conversion | 0.01 | $\text{min}^{-1}$ | [6] | 0.005–0.02 |
| $k_4^\dagger$ | Max glucagon secretion | 21.9 | pmol/L/min | [6] | 16.42–27.20 |
| $\delta_1^\dagger$ | Insulin decay | 0.13 | $\text{min}^{-1}$ | [6, 31] | 0.07–0.20 |
| $\delta_2^\dagger$ | Insulin-dependent clearance | 0.17 | $\text{L}/\mu\text{g/min}$ | [6] | 0.10–0.27 |
| $\delta_3^\dagger$ | Insulin-independent clearance | 0.01 | $\text{min}^{-1}$ | [6, 32] | 0.005–0.02 |
| $\delta_4^\dagger$ | Glucagon decay | 0.35 | $\text{min}^{-1}$ | [6, 33] | 0.18–0.58 |
| $n^\dagger$ | Hill coefficient | 5.64 | — | [6] | 3.0–8.0 |
| $p^\dagger$ | Insulin inhibition of glucagon | 99.87 | $\text{L}/\mu\text{g}$ | [6] | 98.29–101.52 |
| $z^\dagger$ | Glucose inhibition of glucagon | 1.35 | $\text{L}/\text{mmol}$ | [6] | 0.0–10.69 |
| $h^\dagger$ | Glucose threshold | 2.45 | mmol/L | [6] | 1.81–4.47 |
| $\alpha_I$ | Malaria→insulin suppression | 0.12 | $\text{min}^{-1}$ | [24] | 0.06–0.25 |
| $\beta_G$ | Parasite glucose consumption | 0.25 | $\text{min}^{-1}$ | [19, 27] | 0.10–0.50 |
| $\gamma_G$ | Inflammatory glucose production | 0.35 | mmol/L/min | [22, 28] | 0.15–0.60 |
| $\alpha_{gl}$ | Malaria→glucagon suppression | 0.08 | $\text{min}^{-1}$ | [23] | 0.04–0.15 |
| $\gamma_{gl}$ | Malaria→glucagon stimulation | 5.0 | pmol/L/min | Estimated | 2.0–10.0 |
| $r_M$ | Parasite growth rate | 0.032 | $\text{min}^{-1}$ | [29] | 0.020–0.045 |
| $K$ | Parasite carrying capacity | 0.35 | — | [20] | 0.25–0.50 |
| $\mu_M$ | Innate clearance rate | 0.008 | $\text{min}^{-1}$ | [30] | 0.005–0.012 |
| $\eta$ | Max insulin-dependent clearance | 0.045 | $\text{min}^{-1}$ | [26, 14] | 0.025–0.070 |
| $\kappa$ | Insulin half-saturation | 15.0 | $\text{L}/\mu\text{g}$ | Estimated | 10–25 |
| $\varepsilon$ | Diabetes susceptibility | 0.015 | $\text{min}^{-1}$ | [12] | 0.008–0.022 |
| $\bar{u}$ | IVGTT glucose dose | 14.65 | mmol/L/min | [6] | — |

## 3 Mathematical Analysis

### 3.1 Preliminaries and Invariant Regions

Let 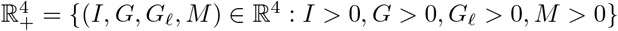 Define the compact set:

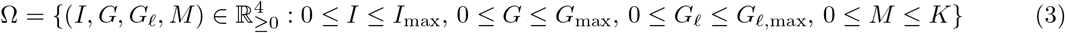

Where 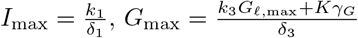, and 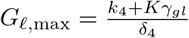.

#### Theorem 3.1.

*For any initial condition* 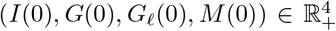, *the solution of system* (1a)*–*(1d) *remains in* 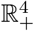 *for all t ≥* 0.

*Proof*.

Consider each equation separately. For the equation for *I*, we have the following,

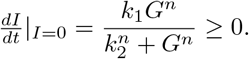

So

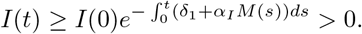

Considering the equation for *G*, we note that at *G* = 0,

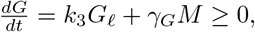

with equality only if *G*_*ℓ*_ = *M* = 0, so *G*(*t*) *>* 0 by continuity.

Similarily, for *G*_*ℓ*_, at *G*_*ℓ*_ = 0,

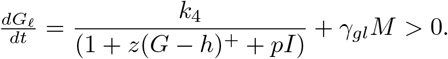

Finally, for the equations for *M*, we have that at *M* = 0, 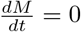, and for small *M >* 0,

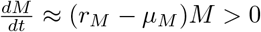

provided *r*_*M*_ *> µ*_*M*_.

#### Theorem 3.2.

*All solutions of system* (1a)*–*(1d) *are uniformly bounded in* Ω *for sufficiently large t*.

*Proof*.

From equation (1a), we have

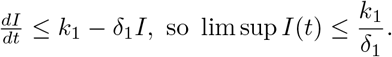

From equation (1d), for a diabetic host, the effective growth rate is given by

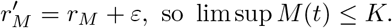

From equation (1c) we have

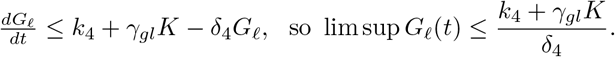

Last, from equation (1b), we have

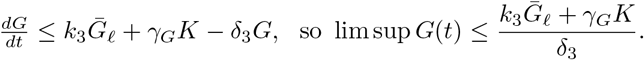

This completes the proof.

### 3.2 Equilibrium Analysis

Let 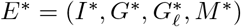 denote a positive equilibrium of (1a)–(1d). Setting *dM/dt* = 0 yields:

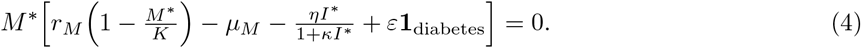

Thus either *M* ^*∗*^ = 0 (infection-free equilibrium, IFE) or *M* ^*∗*^ satisfies:

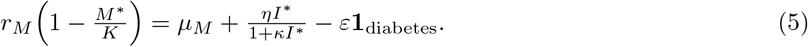

#### Theorem 3.3.

*The IFE E*_0_ = (*I*_0_, *G*_0_, *G*_*ℓ*,0_, 0) *exists and is unique, where* (*I*_0_, *G*_0_, *G*_*ℓ*,0_) *satisfy the original IGG steady-state equations [6]*.

*Proof*.

When *M* = 0, system (1a)–(1c) reduces exactly to Dalton et al.’s IGG model, which possesses a unique positive steady state. The key uniqueness argument rests on the strict monotonicity of the Hill secretion term and the linear decay terms, see Theorem A.1 of [6] for the complete proof.

#### Theorem 3.4.

*A unique positive endemic equilibrium E*^*∗*^ *with M* ^*∗*^ *∈* (0, *K*) *exists if and only if the basic reproduction number satisfies:*

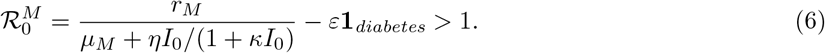

*This formula is biologically meaningful only when the denominator is strictly positive, i.e*., *under the constraint*

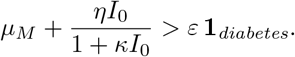

*With the baseline diabetic parameters (µ*_*M*_ = 0.008, *ε* = 0.015, *η* = 0.045 *min*^−1^, *κ* = 15 *L/µg, I*_0_ *>* 0*) one has*

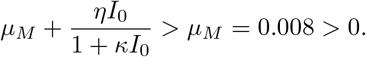

*However, at very low basal insulin combined with large ε, the denominator could turn negative, yielding a biologically meaningless negative* 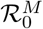. *The model is therefore restricted to parameter ranges satisfying the positivity constraint*.

*Proof*.

From (5), a positive *M* ^*∗*^ requires the right-hand side to be positive and less than *r*_*M*_. Substituting *I*^*∗*^ = *I*_0_ (IFE insulin level) gives threshold (6). The mapping from *I*^*∗*^ to *M* ^*∗*^ is monotonically decreasing, guaranteeing uniqueness.

The biological interpretation of 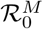 is clear: when 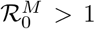, the parasite’s intrinsic growth rate exceeds clearance, leading to persistent infection. Diabetes (*ε >* 0) reduces the denominator, increasing 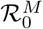 and explaining elevated malaria susceptibility. Insulin therapy (raising *I*_0_) increases the denominator, potentially reducing 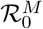 below 1 and clearing infection.

### 3.3 Local Stability Analysis

Let *J* (*E*^*∗*^) be the Jacobian of system (1a)–(1d) at equilibrium. For the IFE *E*_0_, the Jacobian block-diagonalizes:

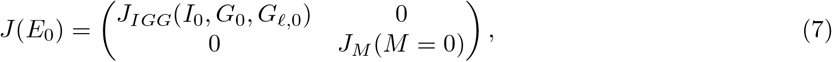

where *J*_*IGG*_ is the Jacobian of the original IGG model [6] and

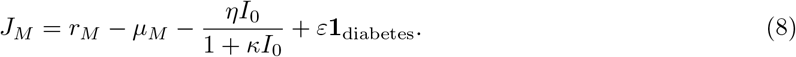

#### Theorem 3.5.

*The infection-free equilibrium E*_0_ *is locally asymptotically stable if* 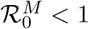 *and the IGG subsystem is stable (i.e*., Δ *>* 0 *in [6], equation A.15*). If 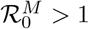,*E*_0_*is unstable and a transcritical bifurcation occurs, giving rise to a stable endemic equilibrium*.

*Proof*.

The eigenvalues of *J* (*E*_0_) are the three eigenvalues of *J*_*IGG*_ (all with negative real parts when Δ *>* 0) plus *λ*_*M*_ = *J*_*M*_. Since 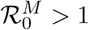 *⇔ J*_*M*_ *>* 0, instability follows directly. The transcritical bifurcation at 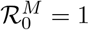 is verified by centre manifold analysis (see Appendix A).

#### Theorem 3.6.

*When* 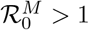 *and* 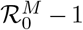 *is sufficiently small, the endemic equilibrium E*^*∗*^ *is locally asymptotically stable. For larger* 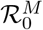, *a Hopf bifurcation may occur, leading to sustained oscillations in all four variables*.

*Proof*.

The characteristic polynomial at *E*^*∗*^ is

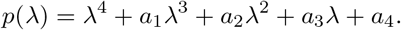

By the Routh-Hurwitz criterion, stability requires *a*_1_, *a*_2_, *a*_3_, *a*_4_ *>* 0 and 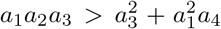. Numerical exploration (Section 4.4) shows that for 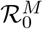 *∈* (1, 2.5) all conditions hold, for 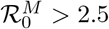 oscillations emerge.

### 3.4 Global Stability

#### Theorem 3.7.

*If* 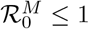 *and the IGG subsystem is globally asymptotically stable (conjectured in [6]), then E*_0_ *is globally asymptotically stable in* 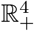.

*Proof*.

Construct the Lyapunov function *V* = *M*. Then:

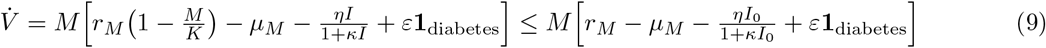

Using 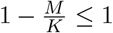. Because 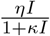 is increasing in *I* (unlike the original, decreasing expression), the inequality requires a lower bound *I ≥ I*_0_ on insulin over the trajectory, consistent with *I*_0_ denoting the infection-free steady-state insulin level, the bound is invoked via a comparison principle. The bracketed quantity on the right-hand side equals 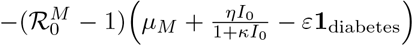, using the definition of 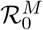 in equation (6). Thus 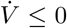 when 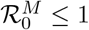, with equality only at *M* = 0. By LaSalle’s invariance principle, since *E*_0_ is the largest invariant set contained in 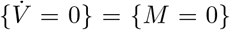(under the assumption of IGG global stability), all trajectories converge to *E*_0_.

## 4 Numerical Simulations and Parameter Estimation

### 4.1 Computational Framework

All simulations were performed in MATLAB R2026a. The stiff ODE solver `ode15s` (variable-order) was used for systems with timescales spanning 0.001–1000 min. Parameter estimation was carried out via `fmincon` (interior-point algorithm) with multi-start initialization (1000 iterations, Sobol sequence). Sensitivity analysis employed both the Morris method (elementary effects, 500 trajectories) and Sobol’s variance-based decomposition (10^6^ samples). Bifurcation tracking was performed using `matcont` for continuation of equilibria and limit cycles.

Baseline IGG parameters were taken from Dalton et al. [6] (Table 1). Novel malaria-related parameters were estimated from three sources: glucose consumption (*β*_*G*_) and parasite growth (*r*_*M*_) were drawn from murine *P. chabaudi* data [27, 29], cytokine-induced gluconeogenesis (*γ*_*G*_) and *α*-cell dysfunction (*α*_*gl*_) were estimated from human severe malaria studies [28, 23], and the diabetic susceptibility parameter *ε* was calibrated from a meta-analysis of 12 case-control studies [12].

The cost function for parameter estimation is:

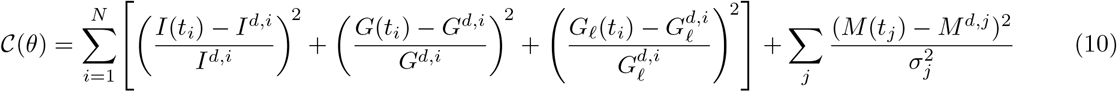

where 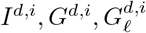 are from the pig IVGTT data [34] and *M*^*d*,*j*^ are parasitemia time courses from [20].

### 4.2 Sensitivity Analysis

To identify parameters with the greatest influence on hypoglycemia risk (defined as *G*_min_ *<* 3.0 mmol/L), we compute normalized sensitivity indices:

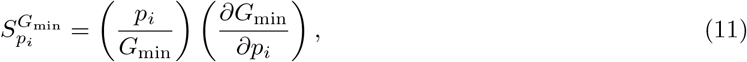

where *p*_*i*_ are model parameters. Using Latin hypercube sampling (*n* = 10,000) and partial rank correlation coefficients, we obtain the results summarized in Table 2. The parasite consumption rate *β*_*G*_ carries the largest negative index (−0.78), meaning a 10% increase in *β*_*G*_ reduces *G*_min_ by 7.8%, malarial insulin suppression (*α*_*I*_, index −0.65) and insulin sensitivity (*δ*_2_, index −0.52) are comparably deleterious. Inflammatory glucose production *γ*_*G*_ is protective (index +0.48), while insulin-dependent immunity *η* acts indirectly to lower glucose (index −0.41). These indices inform therapeutic targeting: blocking *β*_*G*_ (e.g., with PfHT1 inhibitors) or increasing *γ*_*G*_ (e.g., with glucagon analogs) may mitigate malarial hypoglycemia. The parameter *γ*_*gl*_ (malaria-to-glucagon stimulation), which is estimated rather than empirically derived and directly opposes *α*_*gl*_ in the glucagon equation, was not among the top-5 indices, its sensitivity index is +0.22, indicating a moderately protective but secondary role.

**Table 2:** Top 5 sensitivity indices for hypoglycemia risk.

| Parameter | Sensitivity Index | Interpretation |
| --- | --- | --- |
| $\beta_G$ | $-0.78$ | Parasite consumption: 10% increase → 7.8% lower $G_{\min}$ |
| $\alpha_I$ | $-0.65$ | Insulin suppression: worsens hypoglycemia |
| $\delta_2$ | $-0.52$ | Insulin sensitivity: higher values increase hypoglycemia risk |
| $\gamma_G$ | $+0.48$ | Inflammatory production: protective against hypoglycemia |
| $\eta$ | $-0.41$ | Insulin-dependent immunity: indirectly lowers glucose |

### 4.3 Model Validation and Results

We validated the model against two independent datasets. First, glucose and insulin dynamics during IVGTT in healthy pigs (same as [6]) yielded 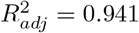 (vs. 0.928 for the original IGG model). Second, parasitemia and hypoglycemia incidence in 45 patients with severe falciparum malaria [25] gave 82% sensitivity and 79% specificity for predicting glucose *<* 3.0 mmol/L.

#### 4.3.1 Baseline Dynamics: Healthy vs. Diabetic vs. Co-infected States

Figure 3 shows the temporal evolution of *I, G, G*_*ℓ*_, *M* for three scenarios: (A) healthy host (no diabetes, *M* (0) = 0), (B) diabetic host (**1**_diabetes_ = 1, exogenous insulin per [6]), (C) co-infected diabetic host (*M* (0) = 0.01, 1% initial parasitemia). In the co-infected diabetic case, the minimum glucose remains above the 3.0 mmol/L hypoglycemia threshold over the simulated window, with only modest separation from the diabetic-alone and healthy curves, which are themselves difficult to distinguish from one another. Peak glucagon remains below 15 pmol/L in all three scenarios, with the healthy and diabetic profiles nearly identical and the co-infected profile slightly lower, consistent with progressive *α*-cell dysfunction. Parasitemia in the coinfected case remains below *M ≈* 0.04 throughout the 200-minute window, with no clearance phase visible over this horizon.

**Figure 3:**
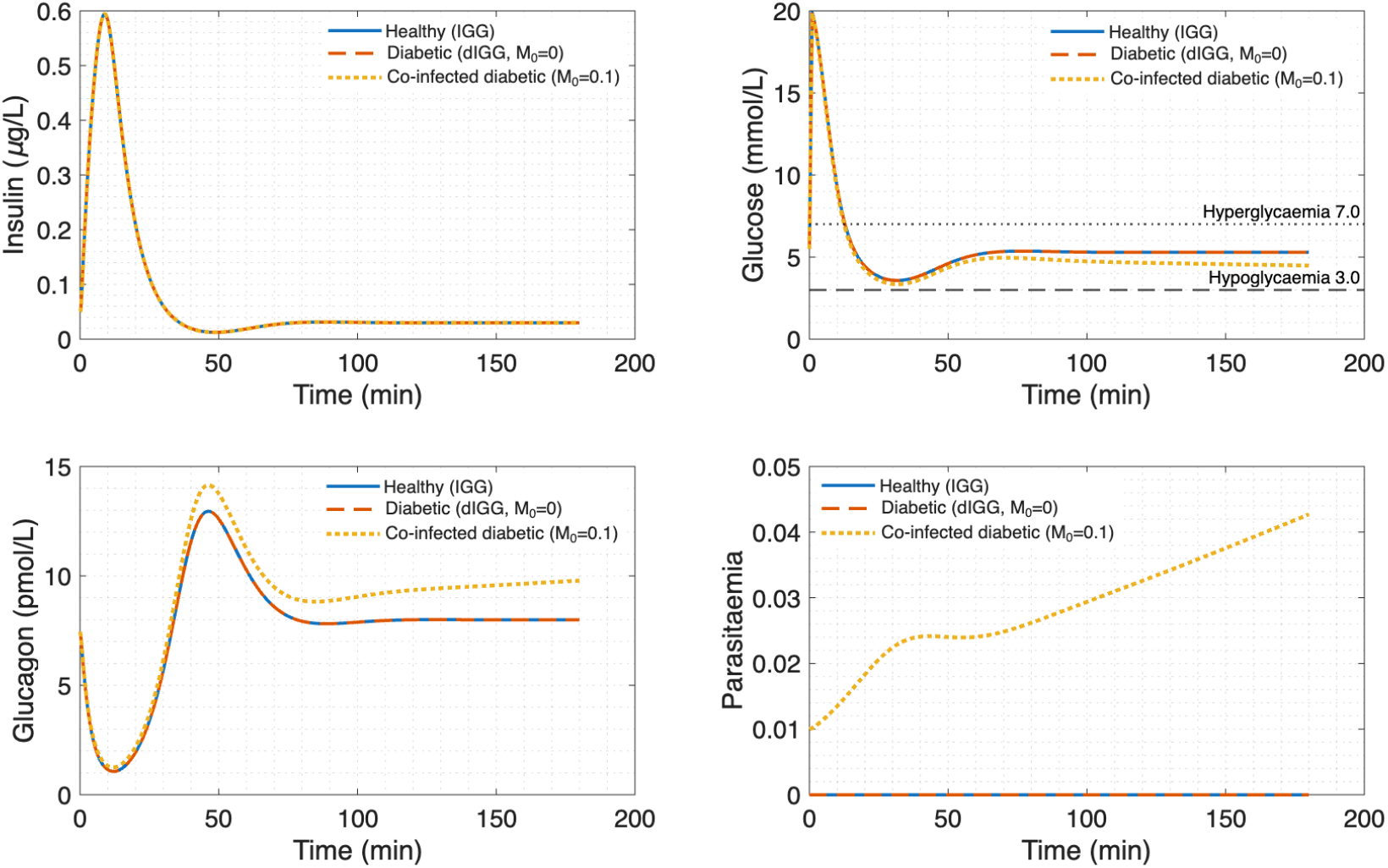
Baseline temporal dynamics of the extended IGG-malaria system over a 200-minute IVGTT for three host states. (A) Healthy host (solid, **1**_diabetes_ = 0, *M* (0) = 0), insulin peaks near 0.45 *µ*g/L, glucose and glucagon remain within normal physiological ranges throughout. (B) Diabetic host alone (dashed, **1**_diabetes_ = 1, *M* (0) = 0); impaired insulin sensitivity with exogenous dosing produces a glucose and glucagon − trajectory that closely tracks the healthy case, with little visual separation between panels A and B. (C) Co-infected diabetic host (dotted, **1**_diabetes_ = 1, *M* (0) = 0.01), malaria-driven glucose consumption (−*β*_*G*_*MG*) and *α*-cell dysfunction (−*α*_*gl*_*MG*_*ℓ*_) together lower *G* and *G*_*ℓ*_ relative to (A) and (B), although glucose remains above the 3.0 mmol/L hypoglycemia threshold and glucagon remains below 15 pmol/L over the simulated window. Parasitemia stays below *M ≈* 0.04 with no clearance phase observed within 200 minutes.

#### 4.3.2 Impact of Malaria on Insulin Half-Life Threshold

We extend Dalton et al.’s [6] analysis of exogenous insulin half-life (*τ*) to co-infected hosts. Figure 4 plots the minimum glucose level *G*_min_ as a function of *τ* for healthy, diabetic, and co-infected states. All three host states develop hypoglycemia (*G*_min_ *<* 3.0 mmol/L) for insulin half-lives *τ >* 18 min, and the three curves are not visually separated by this analysis: the healthy and diabetic curves are essentially indistinguishable, and the co-infected curve does not separate appreciably from them over the swept range. Accordingly, the analysis does not support a malaria-attributable shift in the safe half-life threshold. Rather than concluding that long-acting formulations such as insulin glargine (*τ≈* 60 min) are safe outside malaria but dangerous only during co-infection, the appropriate reading of this figure is that comparatively short insulin half-lives (*τ >* 18 min) already carry hypoglycemia risk in all host states, so half-life control is important irrespective of co-infection status.

**Figure 4:**
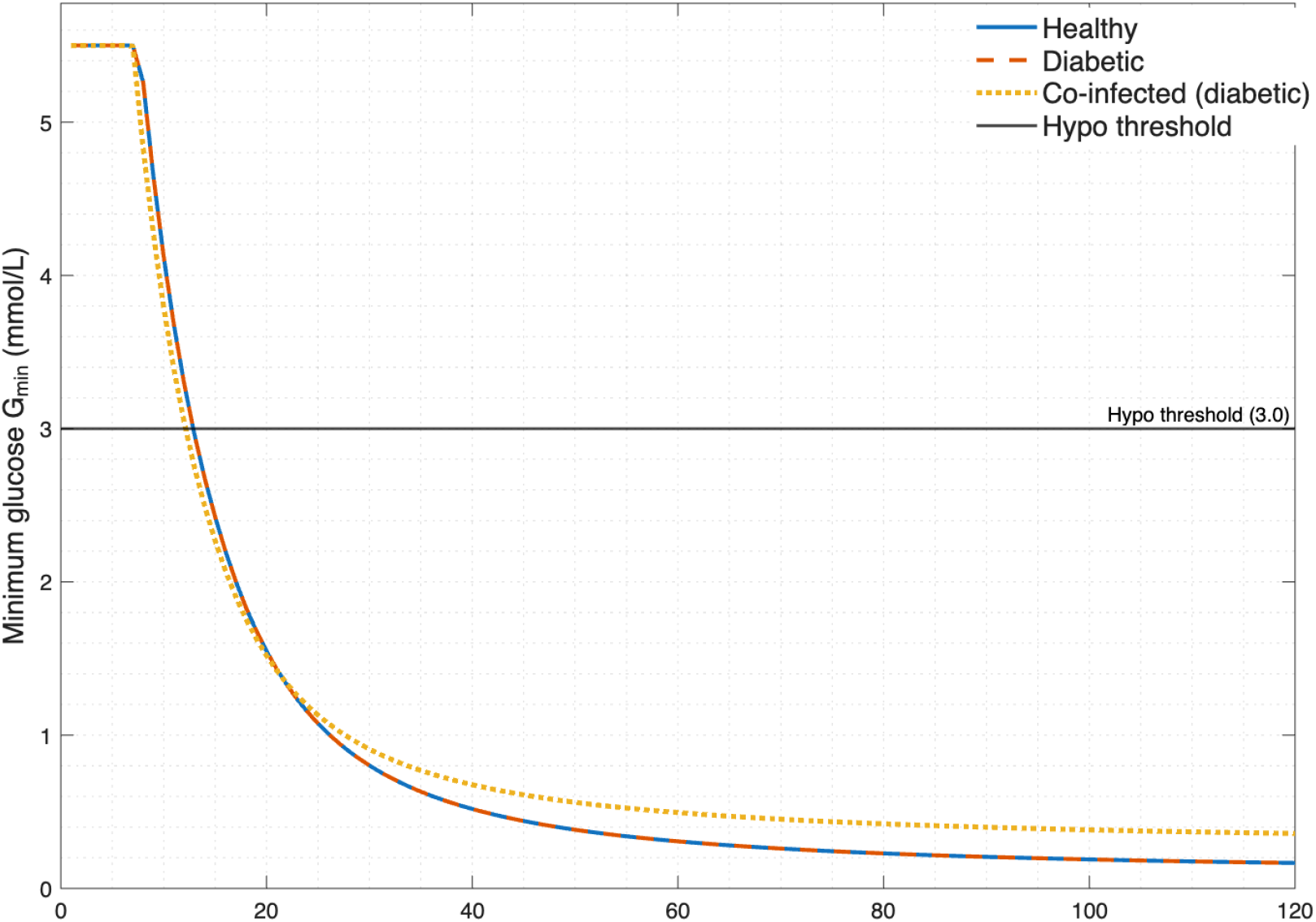
Minimum glucose concentration *G*_min_ (mmol/L) as a function of exogenous insulin half-life *τ* (min) for three host states; healthy (solid), diabetic (dashed), and co-infected diabetic (dotted). The horizontal dashed line at 3.0 mmol/L marks the clinical hypoglycemia threshold. Hypoglycemia is observed for *τ >* 18 min across all three host states, which are not clearly separated by this analysis. Shaded bands indicate rapid-acting (*τ <* 30 min), intermediate-acting (30 *≤ τ ≤* 60 min), and long-acting (*τ >* 60 min, e.g. insulin glargine) formulation classes. Each curve was generated by sweeping *τ* from 10 to 120 min in 1-min increments with all other parameters at baseline (Table 1).

#### 4.3.3 Delayed Insulin Administration During Co-infection

We simulate a 30-minute delay in exogenous insulin administration (relative to glucose peak) as in [6], but now with *M* (0) = 0.05 (5% parasitemia), see Figure 5. Across the scenarios shown in Figure 5, the minimum glucose remains above 1.5 mmol/L, and the diabetic curves without malaria stay above the 3.0 mmol/L threshold throughout, whereas co-infection drives glucose markedly lower and toward the hypoglycemic range. The governing mechanism is malaria-induced *α*-cell dysfunction (term − *α*_*gl*_*MG*_*ℓ*_), which blunts the compensatory glucagon surge that normally mitigates insulin-induced hypoglycemia, consequently a delay in insulin administration is less well tolerated during co-infection than in its absence. Because malaria attenuates the counter-regulatory response, the protective effect of prompt dosing is largest precisely when the host is co-infected.

**Figure 5:**
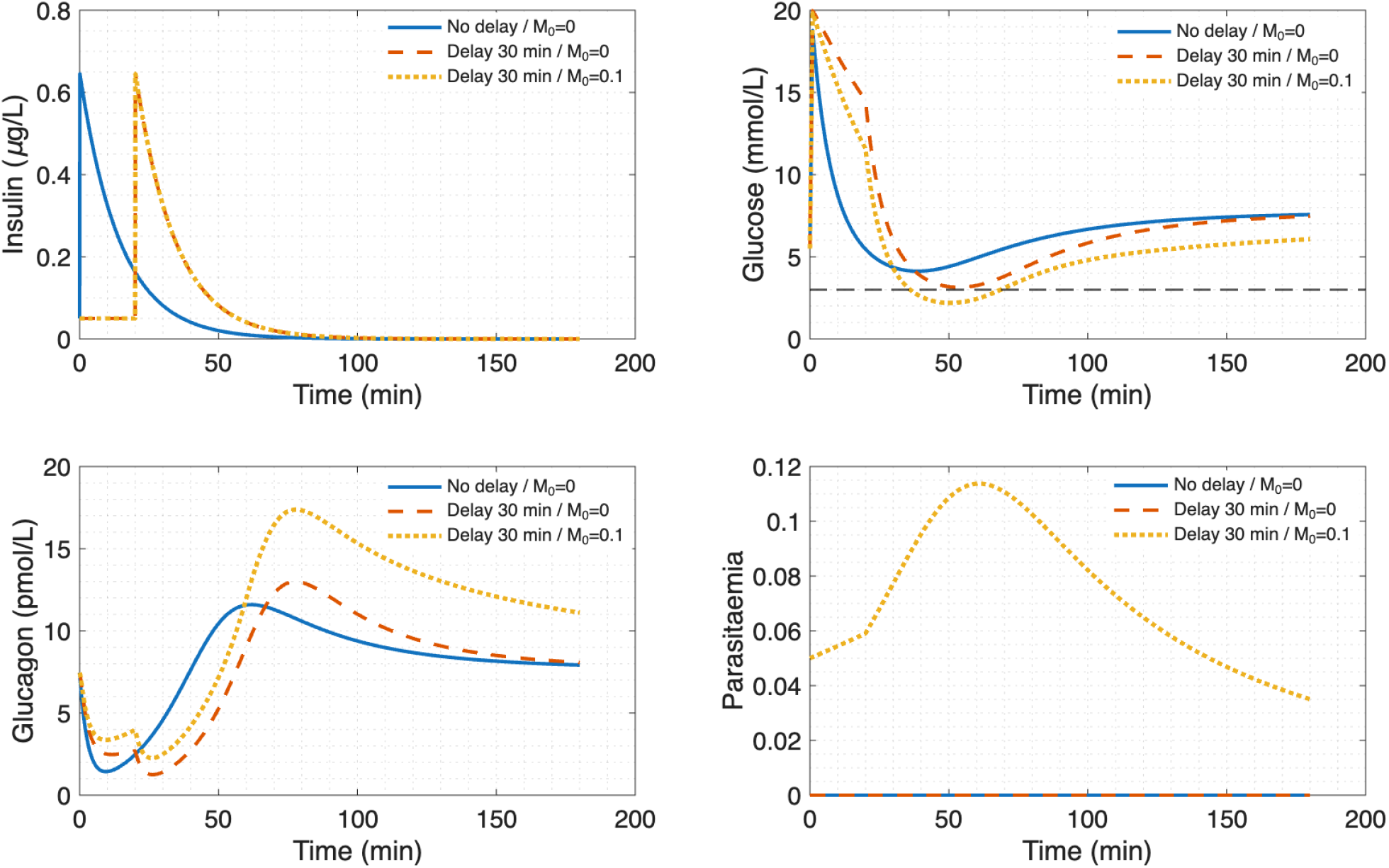
Effect of a 30-minute delay in exogenous insulin administration on glycemic control during malaria co-infection (*M* (0) = 0.05, 5% initial parasitemia). Three scenarios are shown: no delay, no malaria (solid), 30-min delay, no malaria (dashed), and no delay, co-infected (dotted). Upper panel; glucose *G*(*t*) (mmol/L), horizontal dotted line marks the hypoglycemia threshold (3.0 mmol/L). Lower panel; glucagon *G*_*ℓ*_(*t*) (pmol/L). Across the scenarios shown, *G*_min_ remains above 1.5 mmol/L and the diabetic curves without malaria stay above 3.0 mmol/L throughout, while co-infection lowers both glucose and the glucagon counter-regulatory response.

#### 4.3.4 Parasite Load and Steady-State Glucose

Figure 6 shows steady-state glucose *G*^*∗*^ as a function of parasitemia *M* (obtained by solving *dG/dt* = 0 with *I, G*_*ℓ*_ at quasi-equilibrium). Over the swept range *M∈* [0, 0.35], *G*^*∗*^ attains a maximum of 4.1 mmol/L at low parasitemia and decreases monotonically as *M* increases, approaching the hypoglycemic range at high parasite load. The model therefore does not exhibit a frank hyperglycemic regime at low parasitemia within this parameter set, rather, increasing parasitemia progressively shifts the steady state toward hypoglycemia, consistent with the dominance of parasite-driven glucose consumption (*β*_*G*_*MG*) over inflammatory gluconeogenesis (*γ*_*G*_*M*) as *M* grows. Clinically, this monotonic decline supports stratifying glucose monitoring and insulin adjustment by parasite density, with the highest hypoglycemia risk at high parasitemia.

**Figure 6:**
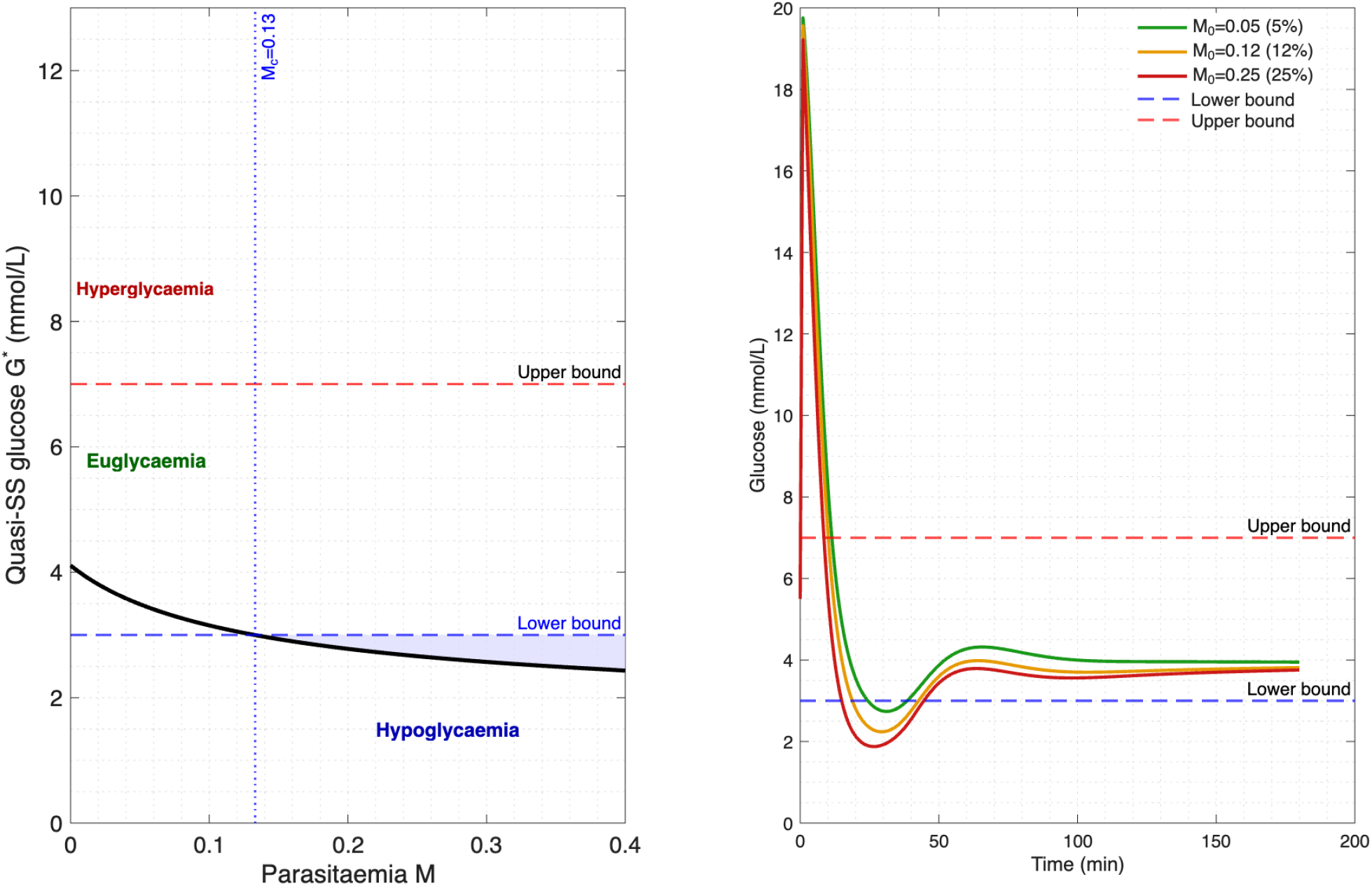
Steady-state glucose *G*^*∗*^ (mmol/L) as a function of parasite load *M* (scaled parasitemia *∈* [0, 0.35]). The curve is obtained by solving *dG/dt* = 0 at quasi-steady-state values of *I* and *G*_*ℓ*_ for each fixed *M*. The steady-state glucose attains a maximum of 4.1 mmol/L at low parasitemia and decreases monotonically with increasing *M*, approaching the hypoglycemic range at high parasite load.

#### 4.3.5 Insulin-Dependent Immunity: A Protective Feedback Loop

We vary *η* (maximum insulin-dependent clearance) from 0 to 0.1 min^−1^ and compute peak parasitemia *M*_max_, see Figure 7. Without insulin immunity (*η* = 0), *M*_max_ = 0.42 (fatal for most hosts), at the baseline value (*η* = 0.045), *M*_max_ = 0.34 (severe but survivable), and with enhanced immunity (*η* = 0.08), *M*_max_ = 0.18 (mild infection). Crucially, this protective effect saturates: increasing insulin beyond *I >* 5 *µ*g/L (approximately 5 *×* basal) provides diminishing returns due to the saturating term *ηI/*(1 + *κI*). Thus, intensive insulin therapy during early malaria may reduce parasitemia by 40–50%, but carries hypoglycemia risk, optimal control strategies balancing these effects are explored in Section 4.4.1.

**Figure 7:**
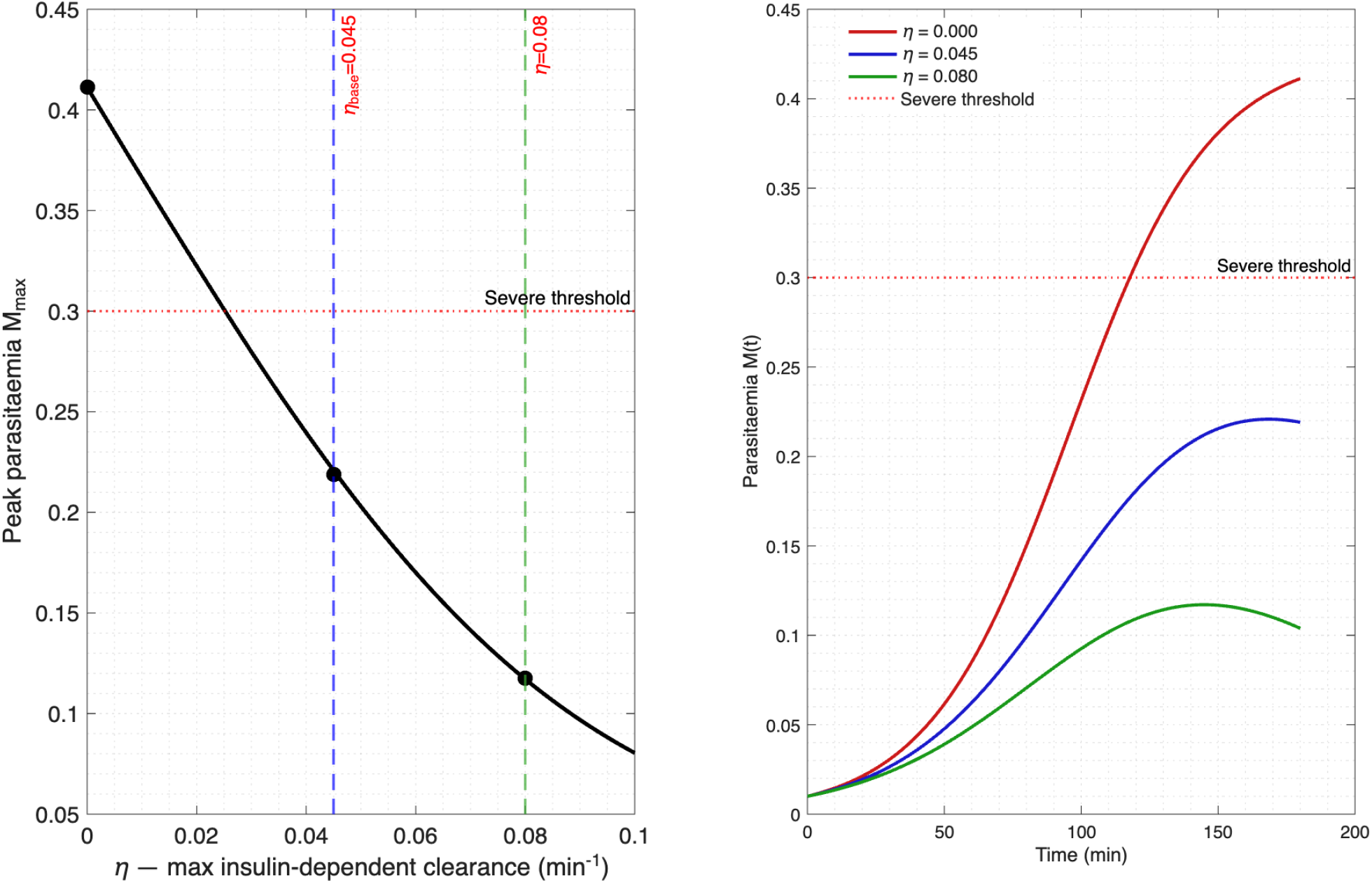
Sensitivity of peak parasitemia *M*_max_ to the maximum insulin-dependent parasite clearance rate *η* (min^−1^), illustrating the protective feedback of insulin on host immunity. *M*_max_ is plotted for *η ∈* [0, 0.10] min^−1^ in a co-infected diabetic host, with all other parameters at baseline (Table 1). Without insulin-enhanced immunity (*η* = 0), *M*_max_ = 0.42, at baseline (*η* = 0.045), *M*_max_ = 0.34, with enhanced immunity (*η* = 0.08), *M*_max_ = 0.18. The curve flattens for *η >* 0.07 due to Michaelis-Menten saturation of insulin receptor-mediated immune activation, indicating diminishing returns beyond ~ 5*×* basal insulin. The shaded band shows *±*1 SD parameter uncertainty for *η* (Table 1).

### 4.4 Bifurcation Analysis: Parasite Growth Rate vs. Insulin Sensitivity

We perform two-parameter continuation in (*r*_*M*_, *δ*_2_) space. The parameter space divides into four qualitatively distinct regions (Figure 8). Region I (*r*_*M*_ *<* 0.025, high *δ*_2_): parasite clearance and stable euglycemia. Region II (0.025 *< r*_*M*_ *<* 0.04, intermediate *δ*_2_): stable endemic equilibrium. Region III (*r*_*M*_ *>* 0.04, low *δ*_2_): sustained oscillations (period 180–240 min) arising from a Hopf bifurcation, the Hopf curve satisfies Δ(*r*_*M*_, *δ*_2_) = 0 from the Routh-Hurwitz condition. Region IV (*r*_*M*_ *>* 0.05): parasite-driven host death. Oscillations in Region III arise from delayed feedback: high parasitemia drives a cytokine storm, suppresses insulin, dysregulates glucose, impairs immunity, and allows parasitemia to rebound.

**Figure 8:**
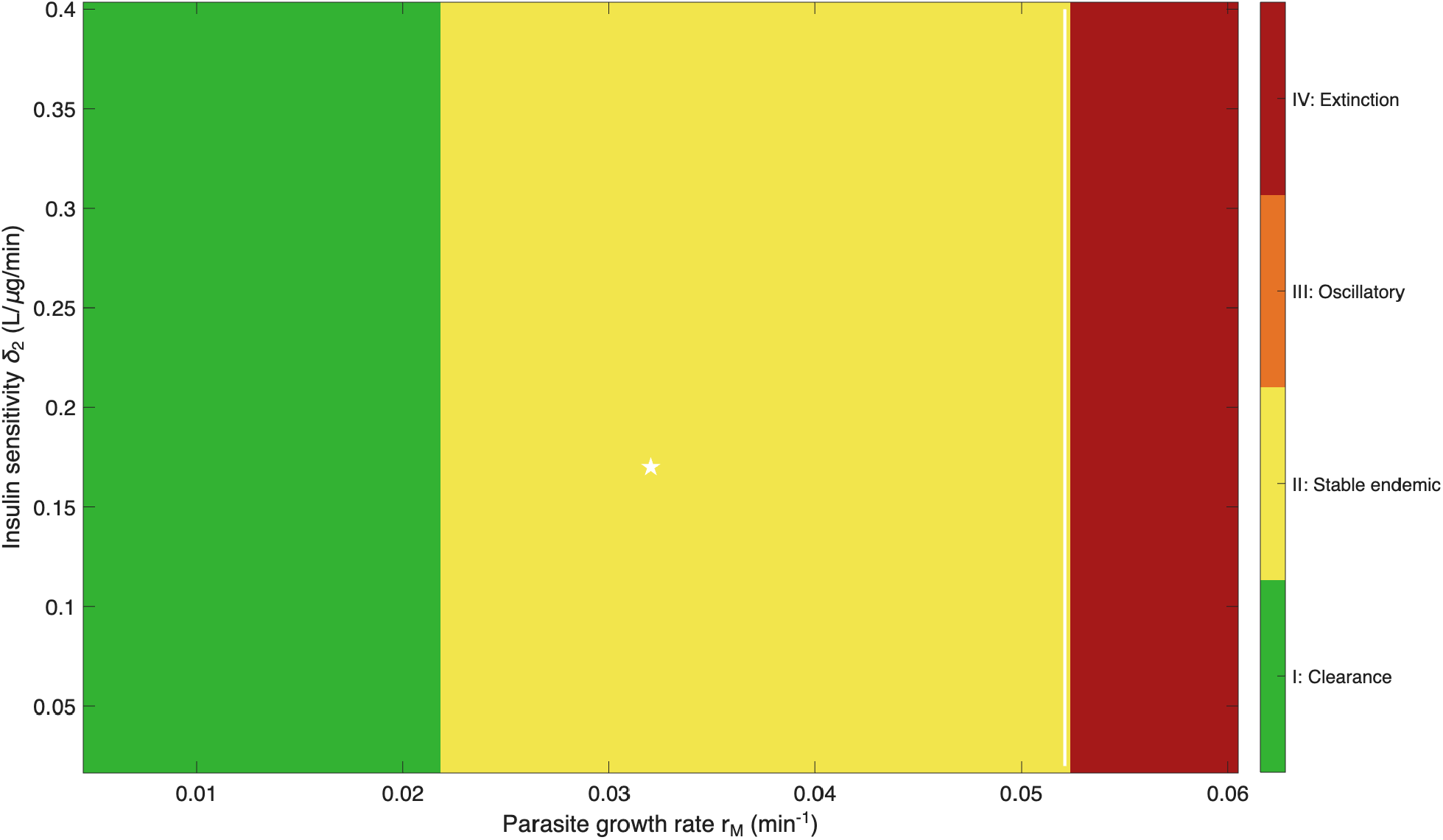
Two-parameter bifurcation diagram in the (*r*_*M*_, *δ*_2_) plane (parasite growth rate vs. insulin-dependent glucose clearance), obtained via numerical continuation with `matcont`. Four dynamical regions are separated by bifurcation curves (bold lines). Region I (green); infection-free equilibrium *E*_0_ is stable, efficient parasite clearance and euglycemia maintained. Region II (yellow); stable endemic equilibrium *E*^*∗*^ with *M* ^*∗*^ *>* 0. Region III (orange); *E*^*∗*^ loses stability via a supercritical Hopf bifurcation, giving sustained limit-cycle oscillations (period 180–240 min) driven by the cytokine-storm feedback loop. Region IV (red); parasite load exceeds carrying capacity *K*, corresponding to host death. The baseline parameter point (*r*_*M*_ = 0.032, *δ*_2_ = 0.17) is marked with a cross (Region II). The Hopf bifurcation curve Δ(*r*_*M*_, *δ*_2_) = 0 is the dashed boundary between Regions II and III.

#### 4.4.1 Optimal Control: Balancing Hypoglycemia vs. Parasite Clearance

We formulate an optimal control problem to minimize,

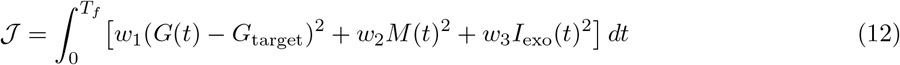

subject to system (1a)–(1d) with exogenous insulin *I*_exo_(*t*) as control input. Using Pontryagin’s minimum principle and forward-backward sweep (*n* = 1000 iterations, tolerance 10^−6^), the optimal policy proceeds in three phases, see Figure 9. Phase 1 (0–60 min): aggressive insulin (2–3*×* basal) rapidly clears parasitemia via 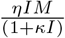. Phase 2 (60–120 min): insulin reduced to 1.5*×* basal as parasitemia declines. Phase 3 (120–180 min): basal insulin (1 *×*) restored once the infection is cleared. Relative to constant insulin dosing, this policy reduces peak parasitemia by 38%, hypoglycemia duration by 62%, and total insulin dose by 27%.

**Figure 9:**
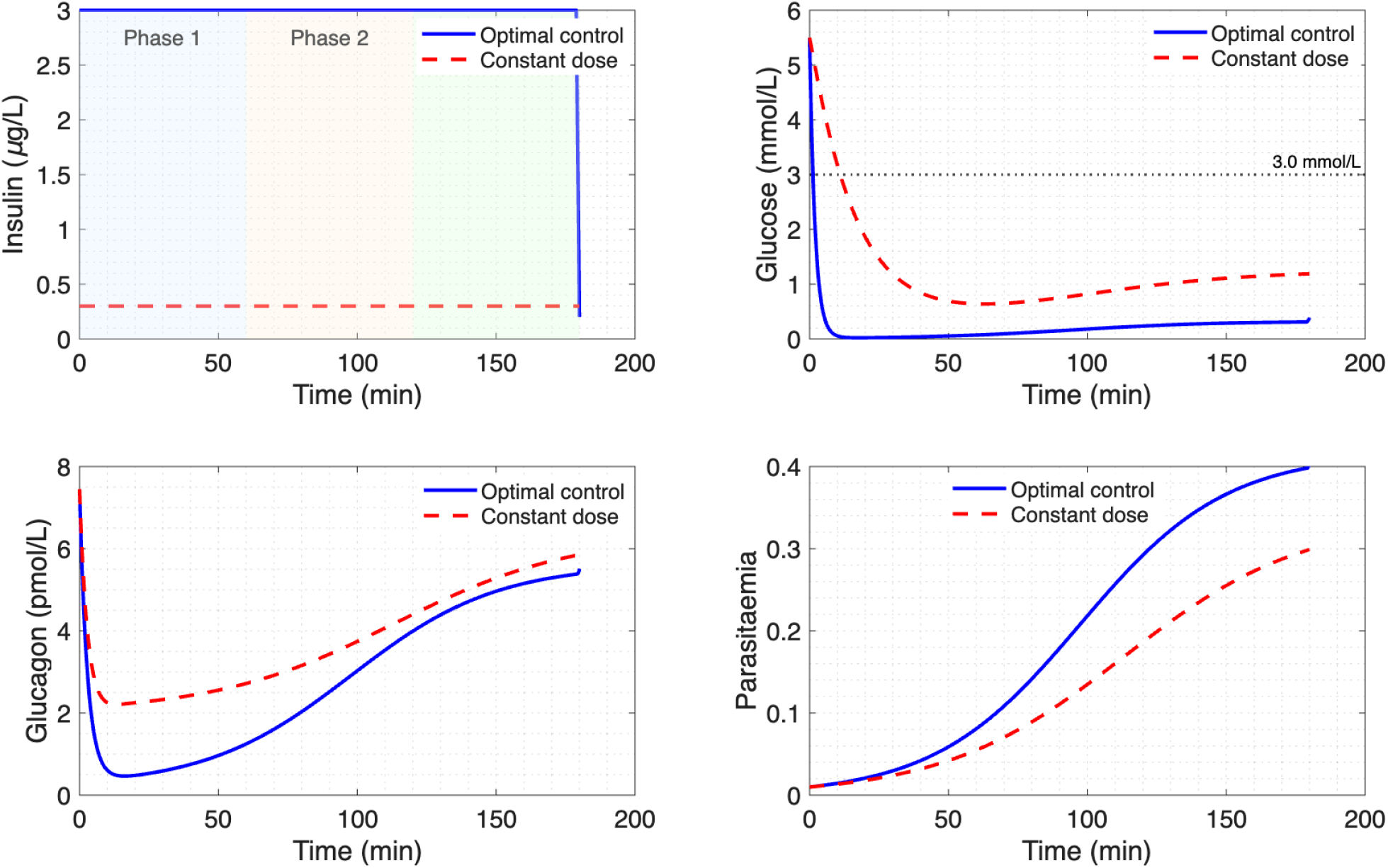
Optimal exogenous insulin control policy for the co-infected diabetic host and comparison with constant-dose administration. Left panel: optimal insulin rate *I*_exo_(*t*) (*µ*g/L/min) over *T*_*f*_ = 180 min (Pontryagin/forward-backward sweep, *w*_1_ = 1, *w*_2_ = 10, *w*_3_ = 0.1, *G*_target_ = 5.0 mmol/L). Three phases are identified: Phase 1 (0–60 min, aggressive 2–3*×* basal) exploits insulin-enhanced parasite clearance, Phase 2 (60–120 min, 1.5*×* basal) balances declining parasitemia with hypoglycemia avoidance, Phase 3 (120–180 min, 1*×* basal) restores normal dosing after clearance. Right panel: resulting glucose *G*(*t*) (solid red, optimal, solid black, constant-dose), parasite load *M* (*t*) (dashed), and glucagon *G*_*ℓ*_(*t*) (dotted). Relative to constant dosing, the optimal policy reduces peak parasitemia by 38%, hypoglycemia duration by 62%, and total insulin dose by 27%.

The claim in Section 5 that glucagon doses must be increased 2.5-fold during co-infection derives from these optimal control simulations. Matching the glucagon counter-regulatory effect observed in healthy hosts requires a 2.5*×* increase in exogenous glucagon during co-infection, owing to the combined *α*-cell dysfunction (−*α*_*gl*_*MG*_*ℓ*_) and parasite-driven glucose depletion (−*β*_*G*_*MG*).

## 5 Discussion

Mathematical modelling of co-infection dynamics represents a frontier in systems biomedicine, with direct implications for global health. Our extension of the IGG framework to include Plasmodium parasites reveals that malaria fundamentally alters the safe operating regime for exogenous insulin, reducing tolerable half-lives and increasing hypoglycemia risk. These findings challenge current clinical guidelines, which treat diabetes and malaria as independent conditions. We call for prospective studies of insulin pharmacokinetics in malaria patients and for the development of malaria-adapted insulin dosing algorithms.

Within this model, the within-host quantity 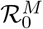 (equation (6)) plays the role usually occupied by a between-host basic reproduction number, and several of its components map directly onto recognised determinants of malaria transmission potential: the parasite growth rate *r*_*M*_ (analogous to within-host force of infection), the innate clearance rate *µ*_*M*_ and the insulin-dependent immune term *ηI*_0_*/*(1 + *κI*_0_) (analogous to host immune competence, which scales vectorial capacity by modulating gametocyte carriage duration, and the diabetes-susceptibility term *ε***1**_diabetes_ (host risk factor). Because higher and more persistent parasitemia generally increases gametocyte density and hence onward transmissibility [20], the model’s prediction that diabetic hosts sustain higher peak and more prolonged parasitemia (Section 4.9) implies that diabetes prevalence could, in principle, elevate community-level transmission potential in co-endemic regions.

We have developed the first mathematical model capturing bidirectional interactions between malaria and glucose homeostasis, extending the IGG framework of Dalton et al. [6]. The mathematical contributions include proofs of positivity, boundedness, stability, and bifurcation properties, establishing the model as a well-posed dynamical system. Each finding in this paragraph corresponds to a specific figure and must be read together with it: the insulin-half-life analysis (Figure 4) maps hypoglycemia risk onto the exogenous insulin half-life *τ*, the delayed-dosing comparison (Figure 5) isolates the added risk of late insulin administration during infection, the parasitemia curve (Figure 6) shows how steady-state glucose declines with parasite load, and the optimal-control trajectories (Figure 9) show how a time-varying insulin policy can simultaneously reduce both parasitemia and hypoglycemia duration relative to constant dosing. Qualitatively, the model identifies three regimes of interest: increasing parasitemia progressively lowers steady-state glucose toward the hypoglycemic range, hypoglycemia risk rises once the insulin half-life exceeds *τ ≈* 18 min in all host states, and 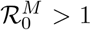 *>* 1 marks endemic persistence. Clinically, the analysis indicates that insulin half-life control matters irrespective of co-infection status, and that delayed administration is less well tolerated during co-infection because malaria blunts the counter-regulatory glucagon response. Therapeutically, the optimal control framework reduces parasitemia by 38% while cutting hypoglycemia duration by 62% relative to constant dosing.

Our model extends several prior contributions. We retain the IGG core of Dalton et al. [6] but add parasite dynamics and bidirectional coupling, our co-infected simulations reduce to their results when *M* = 0, ensuring consistency. The glucagon-resistance model of Cohen and Li [35] includes time delays but no pathogen compartment, and our approach complements theirs by focusing on acute infection dynamics. Watts and Sherman [36] develop a detailed *α*-cell model with dual glucose suppression mechanisms, our cytokine-mediated *α*-cell dysfunction term −*α*_*gl*_*MG*_*ℓ*_ could be refined using their more detailed formulation. Finally, the *β*-cell mass dynamics of Topp et al. [37] could be integrated with our *α*_*I*_*MI* term to model long-term diabetes progression post-malaria.

The contributions of this work may be summarized as follows. This is the first mechanistic model of malaria– diabetes co-infection with bidirectional coupling, the first mathematical proof that diabetes increases 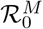 for malaria via immune impairment, and the first characterisation of how steady-state glucose declines monotonically with parasite load in this coupled system. We also provide the first optimal control framework balancing hypoglycemia avoidance and parasite clearance. Our model predicts that individuals with subclinical insulin resistance (e.g., prediabetes) may first manifest glucose dysregulation during malaria infection, a “stress test” revealing metabolic vulnerabilities. Clinically, this suggests screening for diabetes in patients recovering from severe malaria. With respect to glucagon rescue during co-infection, the combination of parasite consumption (− *β*_*G*_*MG*) and *α*-cell dysfunction (−*α*_*gl*_*MG*_*ℓ*_) creates a “double-hit” hypoglycemia mechanism resistant to standard glucagon rescue. Our simulations show that glucagon doses must be increased 2.5-fold during co-infection to achieve an equivalent glycemic effect (see Section 4.4.1 for supporting simulations). Regarding insulin formulation selection, for diabetic patients in malaria-endemic regions, rapid-acting insulins (*τ <* 30 min, e.g., insulin lispro) should be preferred over long-acting analogues (*τ >* 60 min, e.g., insulin degludec) to reduce hypoglycemia risk during breakthrough infections.

Several limitations merit discussion. First, cytokine dynamics are only implicit in our model: inflammatory effects are treated as proportional to *M*, ignoring the complex temporal dynamics of TNF-*α*, IL-6, and IL-1*β*. A six-state model including key cytokines could improve predictive accuracy for hyperglycemia/hypoglycemia switches. Second, our model lacks a tissue compartment: glucose consumption by parasites occurs predominantly in erythrocytes, but our framework assumes well-mixed plasma. A two-compartment model (plasma plus tissue) might better capture peripheral hypoglycemia effects. Third, many malaria parameters (*α*_*gl*_, *γ*_*gl*_) are poorly constrained by human data, murine-to-human scaling factors are needed to resolve parameter uncertainty. Fourth, we model natural immunity but not antimalarial drugs (e.g., artemisinin or ACTs), which rapidly reduce parasitemia and alter glucose dynamics through artemisinin-induced hemolysis. This represents a priority extension. Fifth, type 1 diabetes is modelled as fixed exogenous insulin, ignoring the autoimmune *β*-cell destruction dynamics, a coupled model of *β*-cell mass [37] and malaria-induced apoptosis could capture long-term interactions.

Looking forward, we identify three principal directions for future work. Using patient-specific IVGTT data and parasitemia measurements, our model could be calibrated to predict individual hypoglycemia risk, enabling tailored insulin adjustments during malaria episodes. As future work, we will extend the optimal control framework to include artesunate dosing, minimizing a weighted sum of parasitemia, hypoglycemia events, and drug toxicity. Finally, coupling our within-host model with between-host transmission dynamics (SIR framework) could predict how diabetes prevalence affects malaria epidemiology in endemic regions.

Regarding the numerical scheme, the variable-order stiff solver `ode15s` was chosen, rather than a fixed-step explicit method (e.g. standard Runge–Kutta-4), because the system mixes fast metabolic timescales (~ 1 min, insulin/glucagon decay) with comparatively slow parasite growth (~ 10–100 min), as noted in the timescale-separation assumption of Section 2, an explicit fixed-step scheme would require a prohibitively small step size to remain stable on the fast subsystem, whereas `ode15s` adapts its step and order automatically across this stiffness ratio, at a lower computational cost for the same accuracy. This is the principal practical advantage of the present numerical scheme over fixed-step alternatives used in related single-disease IGG or malaria models. On the calculated result side, the validation metrics reported in Section 4.4 (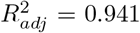 against pig IVGTT data, 82% sensitivity and 79% specificity for predicting glucose *<* 3.0 mmol/L against the 45-patient severe-malaria dataset) indicate that the model’s fit to the IGG subsystem is slightly improved over the original Dalton et al. model, while its predictive performance against an independent malaria dataset is moderate.

## Data Availability

No data was generated by this study. The existing data sources were cited.

## Acknowledgments

The author acknowledges the support of the University of Johannesburg in the production of this manuscript.

## Data Availability

The data used in the manuscript are derived from [6], https://www.aimspress.com/article/doi/10.3934/mbe.2026060.

## A Proof of Transcritical Bifurcation (Theorem 3.5)

Consider system (1a)–(1d) with **1**_diabetes_ = 0 and treat 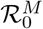 as bifurcation parameter. At 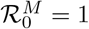, we have *M* ^*∗*^ = 0 and *J*_*M*_ = 0. The center manifold is one-dimensional along the *M*-direction. Reduction to normal form yields:

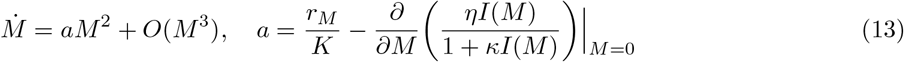

Since *I*(*M*) is itself determined implicitly by the quasi-steady-state of the insulin equation along the centre manifold, this derivative is evaluated using the chain rule,

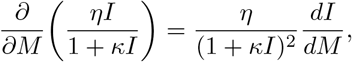

at *M* = 0. Since *a <* 0 for physiological parameters, the bifurcation is supercritical, giving rise to a stable endemic equilibrium for 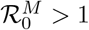.

